# Tumor necrosis factor-α gene polymorphism is associated with short- and long-term kidney allograft outcomes

**DOI:** 10.1101/2021.07.28.21261294

**Authors:** Felix Poppelaars, Mariana Gaya da Costa, Bernardo Faria, Siawosh K. Eskandari, Marc A. Seelen, Jeffrey Damman

**Author notes:** **Address for Correspondence:** Felix Poppelaars, M.D./Ph.D., University Medical Center Groningen, Department of internal medicine, Division of Nephrology, AA53, Postbus 196, 9700 AD Groningen, The Netherlands. Shared second author (Mariana Gaya da Costa, Bernardo Faria).

## Abstract

**Introduction:** Kidney transplantation has excellent short-term results with current immunosuppression regimes, but long-term outcomes have barely improved. Hence, there is a need for new therapeutic options to increase long-term survival of kidney grafts. Drug development for transplantation has slowly plateaued, limiting progress while making drug repurposing an attractive alternative. We, therefore, investigated the impact of tumor necrosis factor-alpha (TNF-α) gene (*TNF*) polymorphisms on kidney graft survival.

**Methods:** We performed a prospective cohort study to assess the association of *TNF* polymorphisms (rs1800629 G>A and rs3093662 A>G) with primary non-function (PNF) and death-censored kidney allograft survival in 1,271 kidney transplant pairs from the University Medical Center Groningen in The Netherlands.

**Results:** The G-allele of the *TNF* rs3093662 polymorphism in donor kidneys was associated with a higher risk of PNF (odds ratio: 2.05; 95%-CI: 1.06-3.97; *P* = 0.032). Furthermore, the G-allele of this *TNF* rs3093662 polymorphism in the donor was also associated with worse 5-year, 10-year, and 15-year death-censored kidney graft survival (*P* < 0.05). The cumulative incidence of graft loss was 15.9% in the reference AA-genotype group and 25.2% in the AG/GG-genotype group, respectively. In multivariable analysis, the association between the *TNF* rs3093662 polymorphism in the donor and 15-year death-censored kidney graft survival remained significant (hazard ratio: 1.51; 95%-CI: 1.05-2.19, *P* = 0.028).

**Conclusion:** Kidney allografts possessing a high-producing *TNF* polymorphism have a greater risk of immediate and late graft loss. Our study adds to a growing body of literature indicating the potential of TNF-α blockade in improving kidney transplantation outcomes.

## Introduction

Kidney transplantation has excellent short-term outcomes with current immunosuppression regimes, while long-term allograft survival has barely improved over the past decades.^1^ Consequently, there is a need for new therapies that can increase the long-term survival of kidney grafts in transplant recipients. Drug development in nephrology and transplantation has been slow and limited in the past twenty years for a multitude of reasons, most notable of which are the steep costs of developing new drugs and identifying and enrolling the patients for clinical trials.^2,3^ Multi-disciplinary efforts are now underway to accelerate the process of drug development for renal transplant patients.^3,4^ Simultaneously, drug repurposing is seen as an attractive alternative to the traditional drug discovery process.^5^ With drug repurposing investigators explore new indications of use for investigational or previously approved drugs. Notably, this approach is low-cost, time-saving, risk-averse, and, therefore, highly efficient.^5^ Recycling existing anti-inflammatory drugs for kidney transplantation, thus, should be considered to improve long-term transplant outcomes.

Cytokines induce and enhance the immune response to combat invading microorganisms, but can also cause severe host injury or disease if the activation of this inflammatory response is excessive or unwanted.^6^ Among all cytokines, tumor necrosis factor-alpha (TNF-α) continues to be one of the most extensively studied.^7^ Currently, there are five therapeutic agents in clinical use that target TNF-α; Four monoclonal antibodies (mAb; Infliximab, Centolizumab, Adalimumab, and Golimumab) and one fusion protein (Etanercept).^8^ The clinical success of TNF-α targeted therapies in rheumatologic disorders and inflammatory bowel diseases has helped to establish these biologicals as the mainstay treatment for autoimmune diseases.^9,10^ Currently, there is substantial evidence that TNF-α plays an equally important role in renal inflammation and fibrosis.^11^ Several studies have also demonstrated the importance of TNF-α in renal ischemia-reperfusion injury by using a variety of TNF-α blocking strategies.^12–14^ On this basis, TNF-α inhibitors have therefore been proposed for use in kidney transplant recipients to improve outcome.^15^

Here, we used human genetics to probe whether targeting TNF-α in renal transplantation will provide clinical benefit. Studies of human genetics are powerful tools in validating therapeutic targets because genetically-backed drug targets are reported to be twice as likely to lead to approved therapeutics compared to traditional drug discovery targets.^16,17^ Hence, we investigated the association between two common functional TNF-α gene (*TNF*) polymorphisms and kidney graft survival in donor-recipient renal transplant pairs.

## Methods

### Patient selection and study end-point

Patients were enrolled if they underwent single kidney transplantation between March 1993 and February 2008 at the University Medical Center Groningen (UMCG) in the Netherlands. A total of 1,430 kidney transplantations were screened for analysis. Of these, 1,271 donor and recipient kidney transplant pairs were included in the study as previously described.^18–20^ Exclusion criteria included lack of DNA, technical complications during surgery, loss of follow-up or re-transplantation. The current work is in line with the declaration of Helsinki and all subjects provided written informed consent. Additionally, the study was approved by the medical ethics committee of the UMCG under file n° METc 2014/077. The primary endpoint of our study was primary non-function (PNF) and long-term death-censored graft survival, defined as the need for re-transplantation or dialysis.

### DNA extraction and *TNF* genotyping

Blood was obtained for the isolation of peripheral blood mononuclear cells or splenocytes were collected from the donors and recipients. Isolation of DNA was performed with a commercial kit according to the manufacturer’s instructions and stored at -80°C. Genotyping of single nucleotide polymorphisms (SNPs) was determined using the Illumina VeraCode GoldenGate Assay kit as per the manufacturer’s instructions (Illumina, San Diego, CA, USA). Genotype clustering and calling were performed using BeadStudio Software (Illumina). The overall genotype success rate was 99.5% and 6 samples with a high missing call rate were excluded from subsequent analyses.

### Statistical analysis

SPSS software version 25 (SPSS Inc, Chicago, IL, USA) was used to perform statistical analyses. Data are presented as median [IQR] for non-parametric variables, mean ± standard deviation for parametric variables, and nominal variables as the total number of patients with percentage [n (%)]. Groups were assessed for differences with the Mann-Whitney U test for not-normally divided variables or Student t-test for normally distributed variables, and χ^2^ test for categorical variables, respectively. The Log-rank test was used to determine the difference in the graft loss frequency between different genotypes. Univariable analysis was used to assess the association of genetic, recipient, donor, and transplant characteristics with graft loss. The variables identified in these analyses were next examined in a multivariable Cox regression with a stepwise forward selection. Statical testing was 2-tailed and *P*<0.05 was regarded as significant.

## Results

### Population characteristics

The characteristics of all donor and recipient kidney transplant pairs are shown in Table 1. The maximum study period was 15 years, with a mean follow-up after transplantation of 6.16 years ± 4.21. During the study period, 214 grafts (17.4%) were lost with the main causes of graft loss being rejection (n = 126), surgical complications (n = 33), recurrence of primary disease (n = 16), vascular causes (n = 12), other causes (n = 16) and unknown (n = 11). By univariable analysis, recipient age, donor age, donor type (living *vs* cadaveric), recipient blood type (AB *vs* others), donor blood type (AB *vs* others), cold ischemia time, warm ischemia time, use of corticosteroids, use of cyclosporin, and delayed graft function (DGF) were found to be associated with graft loss (P < 0.05).

**Table 1:**
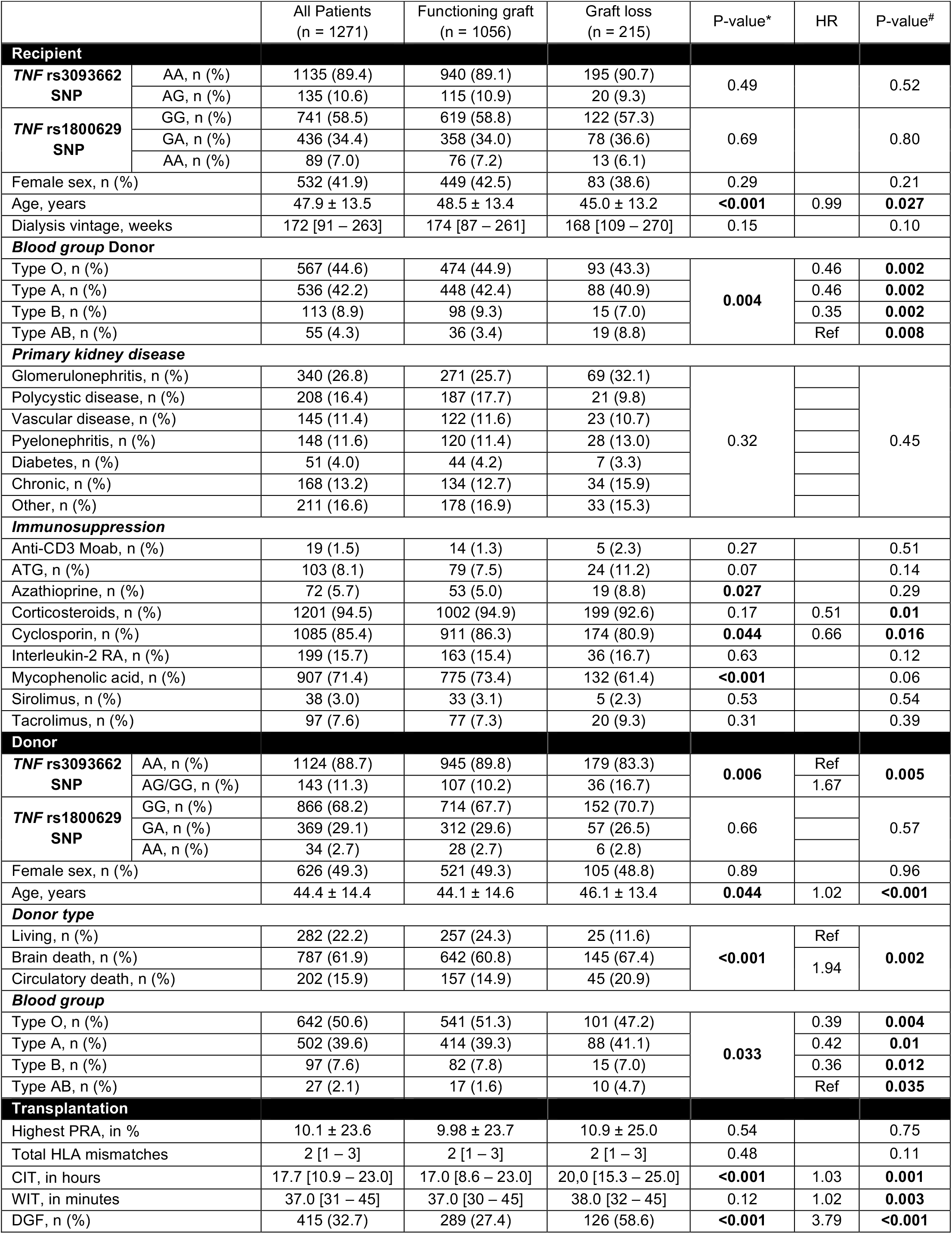
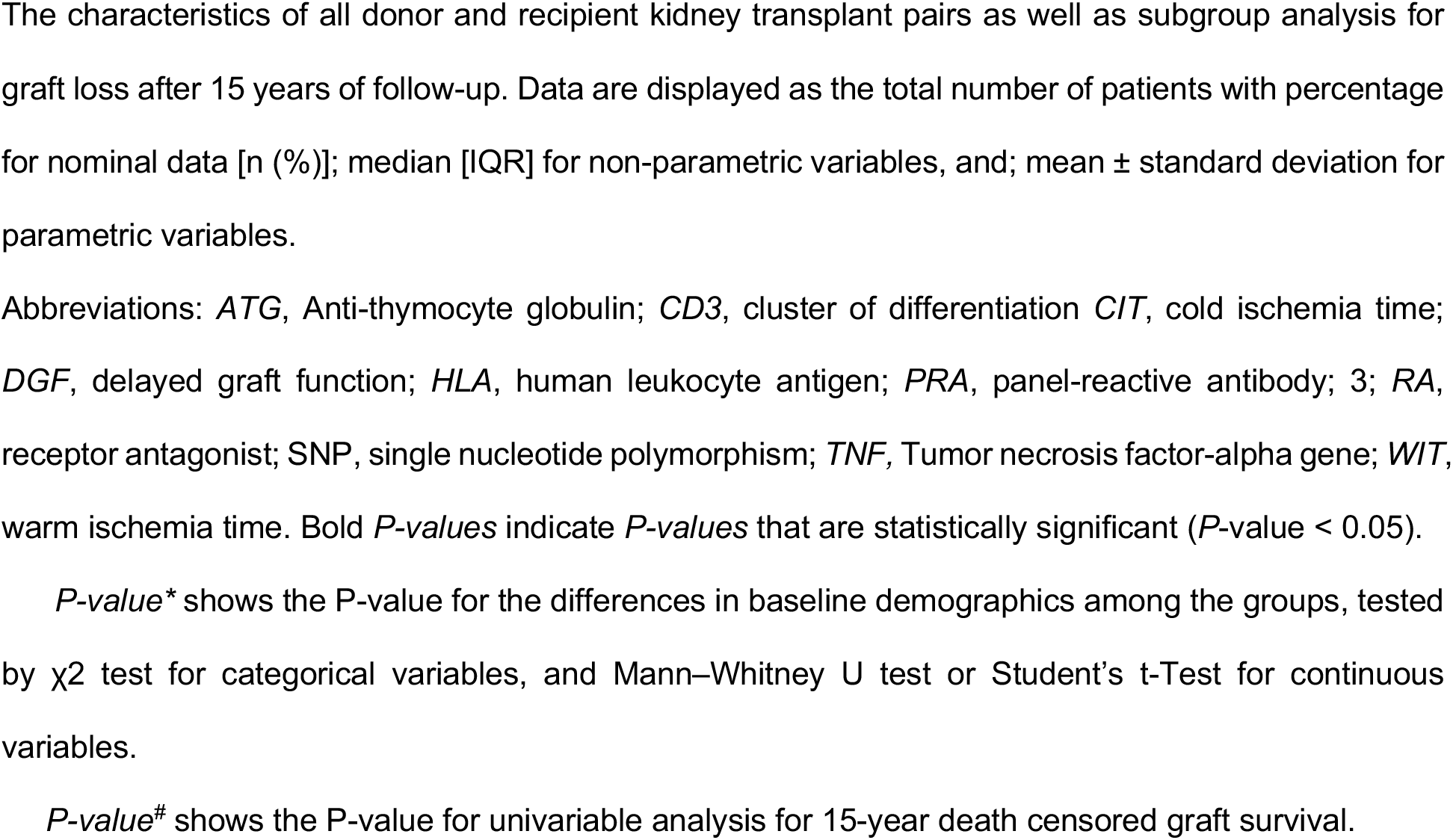
Baseline characteristics of the donors and recipients.

|  |  | All Patients<br>(n = 1271) | Functioning graft<br>(n = 1056) | Graft loss<br>(n = 215) | P-value* | HR | P-value# |
| --- | --- | --- | --- | --- | --- | --- | --- |
| Recipient |  |  |  |  |  |  |  |
| TNF rs3093662<br>SNP | AA, n (%) | 1135 (89.4) | 940 (89.1) | 195 (90.7) | 0.49 |  | 0.52 |
|  | AG, n (%) | 135 (10.6) | 115 (10.9) | 20 (9.3) |  |  |  |
| TNF rs1800629<br>SNP | GG, n (%) | 741 (58.5) | 619 (58.8) | 122 (57.3) | 0.69 |  | 0.80 |
|  | GA, n (%) | 436 (34.4) | 358 (34.0) | 78 (36.6) |  |  |  |
|  | AA, n (%) | 89 (7.0) | 76 (7.2) | 13 (6.1) |  |  |  |
| Female sex, n (%) |  | 532 (41.9) | 449 (42.5) | 83 (38.6) | 0.29 |  | 0.21 |
| Age, years |  | 47.9 ± 13.5 | 48.5 ± 13.4 | 45.0 ± 13.2 | <0.001 | 0.99 | 0.027 |
| Dialysis vintage, weeks |  | 172 [91 – 263] | 174 [87 – 261] | 168 [109 – 270] | 0.15 |  | 0.10 |
| Blood group Donor |  |  |  |  |  |  |  |
| Type O, n (%) |  | 567 (44.6) | 474 (44.9) | 93 (43.3) | 0.004 | 0.46 | 0.002 |
| Type A, n (%) |  | 536 (42.2) | 448 (42.4) | 88 (40.9) |  | 0.46 | 0.002 |
| Type B, n (%) |  | 113 (8.9) | 98 (9.3) | 15 (7.0) |  | 0.35 | 0.002 |
| Type AB, n (%) |  | 55 (4.3) | 36 (3.4) | 19 (8.8) |  | Ref | 0.008 |
| Primary kidney disease |  |  |  |  |  |  |  |
| Glomerulonephritis, n (%) |  | 340 (26.8) | 271 (25.7) | 69 (32.1) | 0.32 |  | 0.45 |
| Polycystic disease, n (%) |  | 208 (16.4) | 187 (17.7) | 21 (9.8) |  |  |  |
| Vascular disease, n (%) |  | 145 (11.4) | 122 (11.6) | 23 (10.7) |  |  |  |
| Pyelonephritis, n (%) |  | 148 (11.6) | 120 (11.4) | 28 (13.0) |  |  |  |
| Diabetes, n (%) |  | 51 (4.0) | 44 (4.2) | 7 (3.3) |  |  |  |
| Chronic, n (%) |  | 168 (13.2) | 134 (12.7) | 34 (15.9) |  |  |  |
| Other, n (%) |  | 211 (16.6) | 178 (16.9) | 33 (15.3) |  |  |  |
| Immunosuppression |  |  |  |  |  |  |  |
| Anti-CD3 Moab, n (%) |  | 19 (1.5) | 14 (1.3) | 5 (2.3) | 0.27 |  | 0.51 |
| ATG, n (%) |  | 103 (8.1) | 79 (7.5) | 24 (11.2) | 0.07 |  | 0.14 |
| Azathioprine, n (%) |  | 72 (5.7) | 53 (5.0) | 19 (8.8) | 0.027 |  | 0.29 |
| Corticosteroids, n (%) |  | 1201 (94.5) | 1002 (94.9) | 199 (92.6) | 0.17 | 0.51 | 0.01 |
| Cyclosporin, n (%) |  | 1085 (85.4) | 911 (86.3) | 174 (80.9) | 0.044 | 0.66 | 0.016 |
| Interleukin-2 RA, n (%) |  | 199 (15.7) | 163 (15.4) | 36 (16.7) | 0.63 |  | 0.12 |
| Mycophenolic acid, n (%) |  | 907 (71.4) | 775 (73.4) | 132 (61.4) | <0.001 |  | 0.06 |
| Sirolimus, n (%) |  | 38 (3.0) | 33 (3.1) | 5 (2.3) | 0.53 |  | 0.54 |
| Tacrolimus, n (%) |  | 97 (7.6) | 77 (7.3) | 20 (9.3) | 0.31 |  | 0.39 |
| Donor |  |  |  |  |  |  |  |
| TNF rs3093662<br>SNP | AA, n (%) | 1124 (88.7) | 945 (89.8) | 179 (83.3) | 0.006 | Ref | 0.005 |
|  | AG/GG, n (%) | 143 (11.3) | 107 (10.2) | 36 (16.7) |  | 1.67 |  |
| TNF rs1800629<br>SNP | GG, n (%) | 866 (68.2) | 714 (67.7) | 152 (70.7) | 0.66 |  | 0.57 |
|  | GA, n (%) | 369 (29.1) | 312 (29.6) | 57 (26.5) |  |  |  |
|  | AA, n (%) | 34 (2.7) | 28 (2.7) | 6 (2.8) |  |  |  |
| Female sex, n (%) |  | 626 (49.3) | 521 (49.3) | 105 (48.8) | 0.89 |  | 0.96 |
| Age, years |  | 44.4 ± 14.4 | 44.1 ± 14.6 | 46.1 ± 13.4 | 0.044 | 1.02 | <0.001 |
| Donor type |  |  |  |  |  |  |  |
| Living, n (%) |  | 282 (22.2) | 257 (24.3) | 25 (11.6) | <0.001 | Ref | 0.002 |
| Brain death, n (%) |  | 787 (61.9) | 642 (60.8) | 145 (67.4) |  | 1.94 |  |
| Circulatory death, n (%) |  | 202 (15.9) | 157 (14.9) | 45 (20.9) |  |  |  |
| Blood group |  |  |  |  |  |  |  |
| Type O, n (%) |  | 642 (50.6) | 541 (51.3) | 101 (47.2) | 0.033 | 0.39 | 0.004 |
| Type A, n (%) |  | 502 (39.6) | 414 (39.3) | 88 (41.1) |  | 0.42 | 0.01 |
| Type B, n (%) |  | 97 (7.6) | 82 (7.8) | 15 (7.0) |  | 0.36 | 0.012 |
| Type AB, n (%) |  | 27 (2.1) | 17 (1.6) | 10 (4.7) |  | Ref | 0.035 |
| Transplantation |  |  |  |  |  |  |  |
| Highest PRA, in % |  | 10.1 ± 23.6 | 9.98 ± 23.7 | 10.9 ± 25.0 | 0.54 |  | 0.75 |
| Total HLA mismatches |  | 2 [1 – 3] | 2 [1 – 3] | 2 [1 – 3] | 0.48 |  | 0.11 |
| CIT, in hours |  | 17.7 [10.9 – 23.0] | 17.0 [8.6 – 23.0] | 20.0 [15.3 – 25.0] | <0.001 | 1.03 | 0.001 |
| WIT, in minutes |  | 37.0 [31 – 45] | 37.0 [30 – 45] | 38.0 [32 – 45] | 0.12 | 1.02 | 0.003 |
| DGF, n (%) |  | 415 (32.7) | 289 (27.4) | 126 (58.6) | <0.001 | 3.79 | <0.001 |

The characteristics of all donor and recipient kidney transplant pairs as well as subgroup analysis for graft loss after 15 years of follow-up. Data are displayed as the total number of patients with percentage for nominal data [n (%)]; median [IQR] for non-parametric variables, and; mean ± standard deviation for parametric variables.
Abbreviations: *ATG*, Anti-thymocyte globulin; *CD3*, cluster of differentiation *CIT*, cold ischemia time; *DGF*, delayed graft function; *HLA*, human leukocyte antigen; *PRA*, panel-reactive antibody; *3*; *RA*, receptor antagonist; *SNP*, single nucleotide polymorphism; *TNF*, Tumor necrosis factor-alpha gene; *WIT*, warm ischemia time. Bold *P-values* indicate *P-values* that are statistically significant (*P-value* < 0.05).
*P-value*\* shows the P-value for the differences in baseline demographics among the groups, tested by $\chi^2$ test for categorical variables, and Mann–Whitney U test or Student's t-Test for continuous variables.
*P-value*# shows the P-value for univariable analysis for 15-year death censored graft survival.

We genotyped two common polymorphisms of the TNF-α gene: rs1800629 G>A in the promoter region at position ∼308 and rs3093662 A>G in the first intron at position ∼851 (Figure 1). The observed genotypic frequencies of the *TNF* SNP rs1800629 G>A were significantly different between recipients (n = 1266; GG, 58.3%; GA, 34.3%; AA, 7.0%) and donors (n = 1269; GG, 68.1%; GA, 29.0%; AA, 2.7%) (*P* < 0.001). More specifically, the A-allele of the *TNF* rs1800629 SNP was more prevalent in kidney transplant recipients. The frequency of the *TNF* rs1800629-A variant were also significantly higher in both donors and recipients than those reported by the 1000 genomes project (*P* < 0.001).^21^ The observed genotypic frequencies of the *TNF* rs3093662 A>G was not significantly different between recipients (n = 1270; AA, 89.3%; GA, 10.6%) and donors (n = 1267; AA, 88.4%; AG, 10.9%; GG, 0.4%) (*P* = 0.08). Yet, the frequency of the *TNF* rs3093662-G variant described in the 1000 genomes project was significantly higher than those in the donors (*P* = 0.003) and recipients (*P* < 0.001).^21^ Due to the low frequency in donors of the *TNF* rs3093662 polymorphism GG-genotype, heterozygotes (AG) and homozygotes (GG)-genotypes were combined to one group (AG/GG). Lastly, the distribution of both polymorphisms was in Hardy-Weinberg equilibrium.

**Figure 1.**
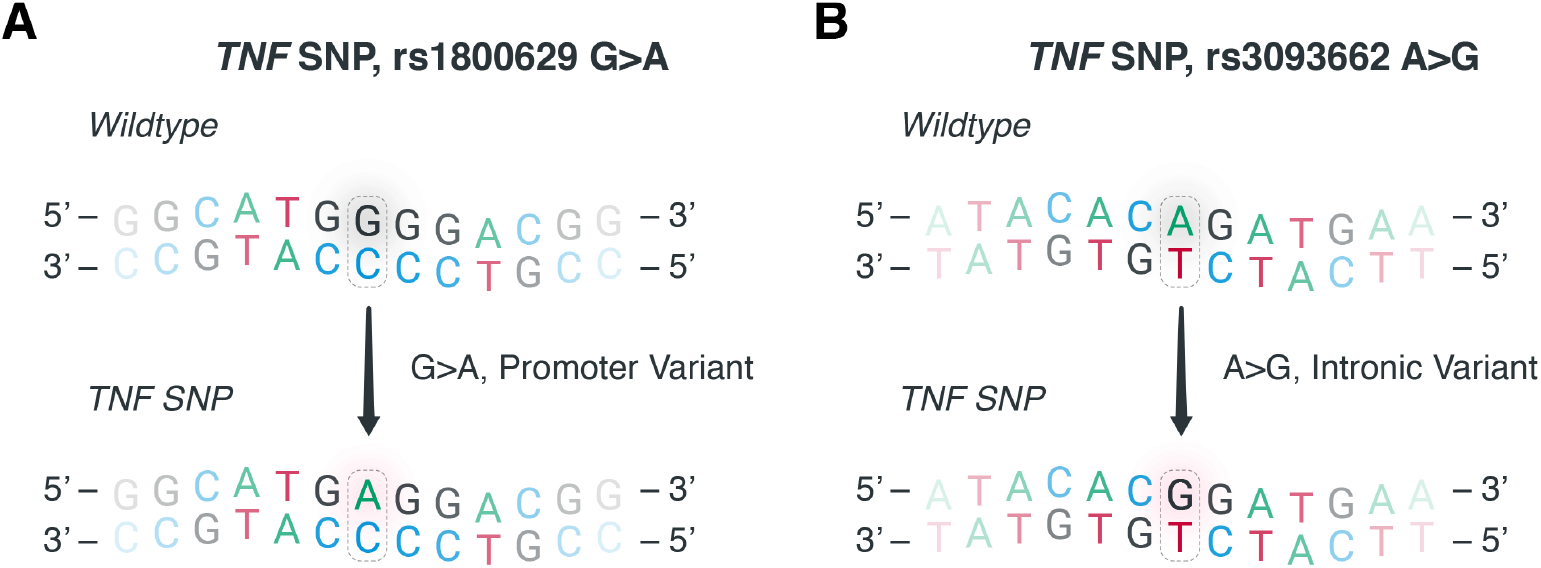
Examined polymorphisms in the tumor necrosis factor-alpha gene. To assess the impact of single nucleotide polymorphisms (SNP) in the tumor necrosis factor-alpha (TNF-α) gene (*TNF*) in kidney transplantation, we assessed the association between graft survival outcomes and the (**A**) *TNF* SNP rs1800629 G>A and the (**B**) *TNF* SNP rs3093662 A>G in the donors and recipients. The *TNF* SNP rs1800629 G>A is located in the promoter region while the *TNF* SNP rs3093662 A>G is in the first intron of the gene.

### A genetic variant in *TNF* is a risk factor for early graft loss after kidney transplantation

We first studied whether genetic variants in *TNF* are associated with PNF to assess the impact on early graft loss. For the *TNF* rs3093662 A>G polymorphism, the proportion of grafts with PNF significantly differed based on the donor genotype (4.3% PNF in the AA-genotype group vs. 8.4% PNF in the AG/GG-group, *P* = 0.029), but not for recipient genotype (*P* = 0.31) (Table 2). In univariable logistic regression, donors carrying the G-allele of the *TNF* rs3093662 A>G polymorphism had a significantly higher risk of PNF (OR = 2.05 compared to A-allele; 95%-CI: 1.06 – 3.97; *P* = 0.032). Next, we performed a multivariable logistic regression analysis with a stepwise forward selection procedure using all characteristics that were significantly associated with PNF in univariable logistic regression (Table 3). In the final model, the *TNF* rs3093662 A>G polymorphism in the donor, donor age, donor type (living *vs* cadaveric), recipient sex, recipient blood type (AB *vs* others), and warm ischemia time were included. After adjustment, the G-allele of rs3093662 in the donor was significantly associated with graft loss with an odds ratio of 2.16 (95% CI: 1.07 – 4.37, *P* = 0.032). For the other *TNF* polymorphism, namely rs1800629 G>A, the proportion of grafts with PNF did not significantly differ based on the recipient genotype (*P* = 0.40), or donor genotype (*P* = 0.35). In conclusion, our data show that the minor allele of the *TNF* rs3093662 A>G SNP in the donor associates with a higher risk of immediate graft loss after kidney transplantation.

**Table 2.** Genotype frequency and hazard ratios for primary non-function based on TNF genotypes.

| *TNF*<br>*rs3093662*<br>*SNP* | Primary non-function |  | Immediate graft function |  | *P*-value | Odds ratio<br>(95%-CI) | *P*-value<sup>b</sup> |
| --- | --- | --- | --- | --- | --- | --- | --- |
|  | AA | AG / GG | AA | AG / GG |  |  |  |
| *Donor* | 48<br>(80.0%) | 12<br>(20.0%) | 1076<br>(89.1%) | 131<br>(10.9%) | **0.029** | **2.05**<br>(1.06 – 3.97) | **0.032** |
| *Recipient* | 56<br>(93.3%) | 4<br>(6.7%) | 1079<br>(89.2%) | 56<br>(4.9%) | 0.31 |  | 0.32 |

Differences in the occurrence of primary non-function (PNF) based on donor and recipient tumor necrosis factor-α genotypes.
Abbreviations: 95-CI%, 95% confidence interval; SNP, single nucleotide polymorphism; *TNF*, recipient tumor necrosis factor-α gene.
Bold *P*-values indicate *P*-values that are statistically significant (*P*-value < 0.05).
<sup>a</sup> *P*-value for the Pearson Chi-square test for differences in genotype frequency.
<sup>b</sup> *P*-value for univariate logistic regression analysis for differences in the incidence of primary non-function.

**Table 3.** Associations of *TNF* polymorphism in the donor with immediate graft loss after kidney transplantation.

| Variables not in the equation |  | Variables in the equation |  |  |
| --- | --- | --- | --- | --- |
| Variables | P-value | Variables | P-value | Hazard Ratio |
| Cold ischemia time<br>(*In hours*) | 0.39 | rs3093662-G in the donor<br>(G *versus* A) | 0.032 | 2.16 (1.07 – 4.37) |
|  |  | Donor age<br>(*In years*) | 0.017 | 1.03 (1.00 – 1.05) |
|  |  | Donor type<br>(*Deceased versus living*) | 0.002 | 9.53 (2.29 – 39.8) |
|  |  | Recipient sex<br>(*Male versus female*) | 0.028 | 0.51 (0.28 – 0.93) |
|  |  | Recipient blood type<br>(*AB versus other*) | 0.027 |  |
|  |  | Warm ischemia time<br>(*In minutes*) | 0.006 | 1.03 (1.01 – 1.05) |

Multivariable logistic regression was performed for primary non-function (PNF) with a stepwise forward selection. All variables with a *P*-value < 0.05 in the univariate analysis for PNF were included. Data are presented as odds ratio with 95%-confidence interval (95%-CI) and *P*-value. In the final model, the *TNF* rs3093662 A>G polymorphism in the donor, donor type, recipient sex, recipient blood type, and warm ischemia time were included, whereas cold ischemia time was not.

### A genetic variant in *TNF* is a risk factor for late graft loss after kidney transplantation

Next, we studied whether the genetic variants in *TNF* are associated with kidney allograft survival to evaluate the relationship between these polymorphisms and late graft loss. No association was found between 15-year death-censored kidney graft survival and the *TNF* rs1800629 G>A polymorphism in the donor (Supplementary data, *P* = 0.64) or the recipient (Supplementary data, *P* = 0.66). Conversely, Kaplan–Meier survival analysis once again revealed that the G-allele of the *TNF* rs3093662 polymorphism in the donor was significantly associated with worse 5-year, 10-year, and 15-year death-censored kidney graft survival (Fig. 2A–C). After 15 years of follow-up, the cumulative incidence of graft loss was 15.9% in the reference AA-genotype group and 25.2% in the AG/GG-genotype group, respectively. The association of the G-allele with late graft loss was maintained when PNF cases were excluded (Fig. 2D, *P* = 0.047), thereby demonstrating that the association between the *TNF* rs3093662 polymorphism and long-term graft survival is independent of its association with early graft loss. We next performed a subgroup analysis for the donor type, since deceased organ donors have elevated levels of circulating pro-inflammatory cytokines. Kaplan-Meier curves demonstrated that the association remained significant between the *TNF* rs3093662 polymorphism and long-term graft survival in kidney allografts from living donors (Fig. 3A, *P* = 0.027) as well as from deceased donors (Fig. 3B, *P* = 0.023). The *TNF* rs3093662 polymorphism in the recipient was not associated with graft survival rates (Supplementary data, *P* = 0.52).

**Figure 2.**
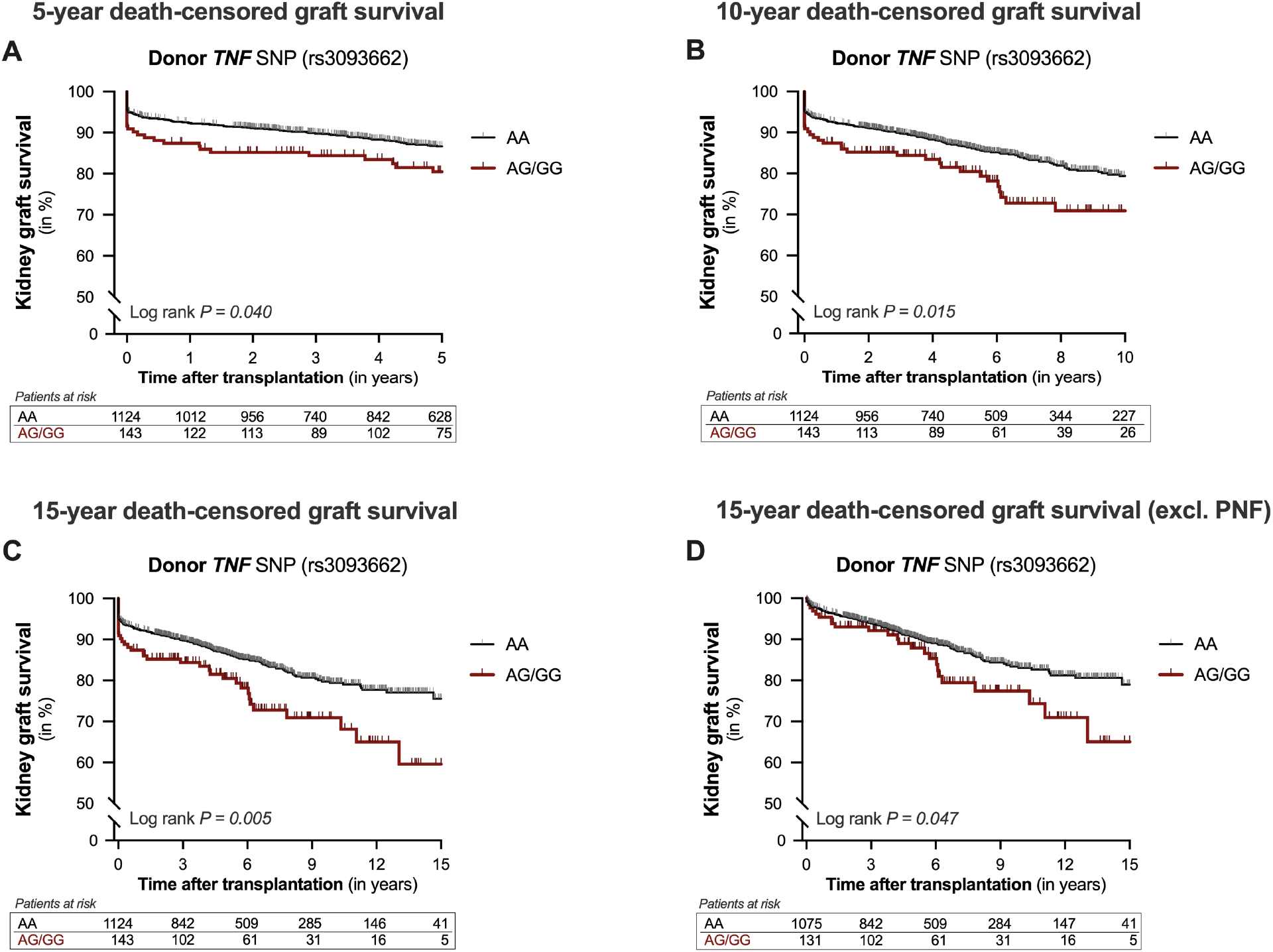
Kaplan-Meier curves for 5-year, 10-year, and 15-year death-censored graft survival after kidney transplantation according to the *TNF* rs3093662 polymorphism in the donor. Cumulative (**A**) 5-year, (**B**) 10-year, and (**C)** 15-year death-censored kidney graft survival according to the presence of the rs3093662 A>G polymorphism in the tumor necrosis factor-alpha gene (*TNF*) in the donor. Furthermore, Kaplan-Meier curves for 15-year death-censored graft survival after kidney transplantation were reanalyzed after excluding donor and recipient kidney transplant pairs with primary non-function (PNF) (**D**). The incidence of graft loss among the groups was compared using the Log-rank test.

**Figure 3.**
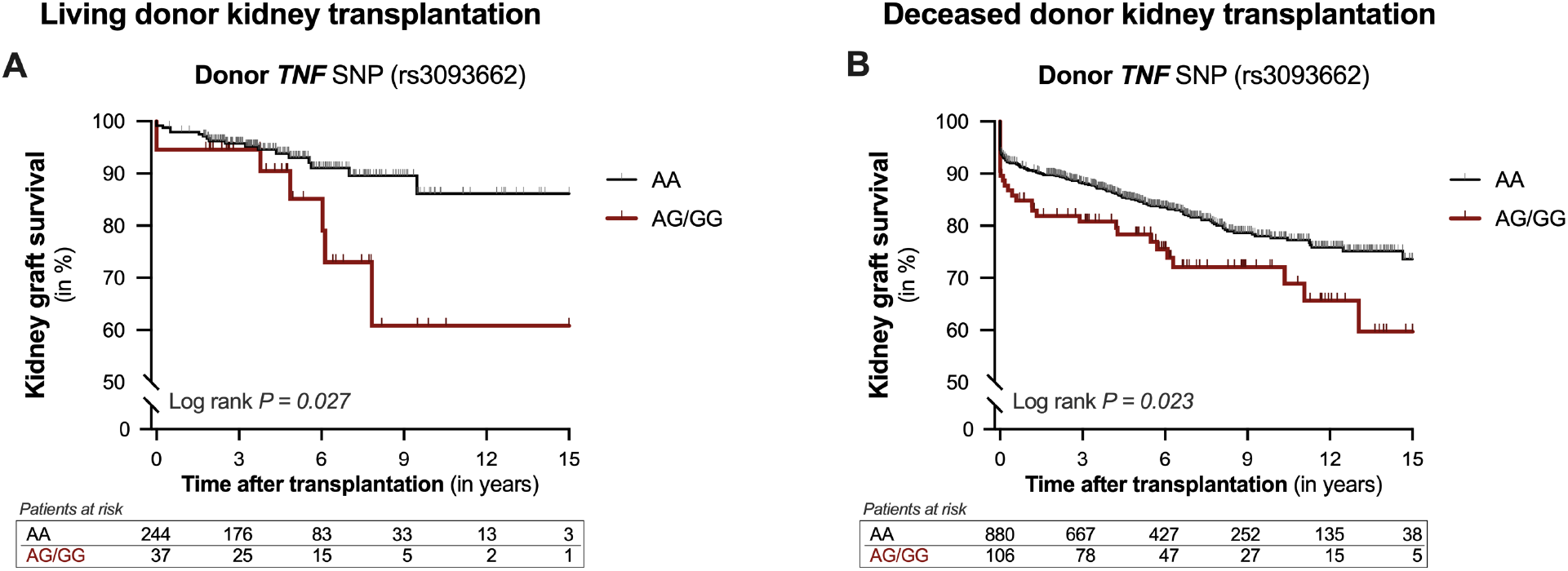
Kaplan-Meier curves for 15-year death-censored graft survival after kidney transplantation according to the *TNF* rs3093662 polymorphism in living and deceased kidney donors. A subgroup analysis for donor type was performed to look at the cumulative 15-year death-censored kidney graft survival according to the presence of the rs3093662 A>G polymorphism in the tumor necrosis factor-alpha gene (*TNF*) in (**A**) living kidney donors and (**B**) deceased kidney donors. The Log-rank test was used to compare the graft loss incidence between the different groups.

We additionally examined whether the G-allele of the *TNF* rs3093662 polymorphism in the donor was independently associated with long-term graft survival. In univariable analysis, the G-allele of the *TNF* rs3093662 polymorphism in the donor was associated with a hazard ratio of 1.67 (95%-CI: 1.16 – 2.38; *P* = 0.005) for graft loss after 15 years of follow-up. Subsequently, we performed a multivariable analysis to adjust for potential confounders using a stepwise forward selection procedure with all clinical variables that were significantly associated with kidney graft survival in univariable analysis (Table 4). In the final model, the *TNF* rs3093662 SNP in the donor, recipient and donor age, recipient blood type (AB *vs* others), and DGF were included. After adjustment, the G-allele of the *TNF* rs3093662 polymorphism in the donor remained significantly associated with graft loss after 15-years with a hazard ratio of 1.51 (95% CI: 1.05 – 2.19, *P* = 0.028). In conclusion, our results demonstrate that the minor allele of the *TNF* rs3093662 (A>G) variant in the donor associates with a higher risk of late graft loss after kidney transplantation.

**Table 4.** Competitive analysis of the associations of characteristics with late graft loss after kidney transplantation.

| Variables not in the equation |  | Variables in the equation |  |  |
| --- | --- | --- | --- | --- |
| Variables | P-value | Variables | P-value | Hazard Ratio |
| Donor blood type<br>(*AB versus other*) | 0.93 | rs3093662-G in the donor<br>(*A versus G*) | 0.028 | 1.51 (1.05 – 2.19) |
| Donor type<br>(*living versus deceased*) | 0.11 | Donor age<br>(*in years*) | 0.002 | 1.02 (1.01 – 1.03) |
| Cold ischemia time<br>(*in hours*) | 0.11 | Recipient age<br>(*in years*) | <0.001 | 0.98 (0.97 – 0.99) |
| Warm ischemia time<br>(*in minutes*) | 0.08 | Recipient blood type<br>(*AB versus other*) | 0.001 |  |
| Corticosteroids | 0.08 | Delayed graft function<br>(*yes versus no*) | <0.001 | 4.00 (3.03 – 5.30) |
| Cyclosporin | 0.27 |  |  |  |

Multivariable cox regression was performed for 15-year death-censored kidney graft survival with a stepwise forward selection. All variables with a *P*-value < 0.05 in the univariate analysis for late graft loss were included. Data are presented as hazard ratio with 95% confidence interval (CI) and *P*-value. In the final model, the *TNF* rs1800629 G>A polymorphism in the donor, recipient and donor age, recipient blood type and delayed graft function (DGF) were included, whereas donor blood type, donor type, cold ischemia time, warm ischemia time, and use of cyclosporin and corticosteroids were not.

## Discussion

The primary finding of our study is that a donor genetic variant in *TNF* associates with an increased risk for graft loss after kidney transplantation. This *TNF* polymorphism in the donor was independently associated with both a higher risk for immediate as well as late graft loss. Extending these findings, the relationship between the *TNF* rs3093662 polymorphism and long-term kidney allograft survival remained significant in a subgroup analysis for different types of kidney transplants. In contrast, long-term graft survival was not impacted by the *TNF* rs3093662 polymorphism in the recipient, nor by the *TNF* rs1800629 polymorphism. In conclusion, our data provide genetic evidence that targeting TNF-α in renal transplantation could be favorable for long-term graft survival in kidney transplantation and encourages the study of TNF-α inhibitors in kidney transplantation in randomized controlled trials.

To our knowledge, our study is the first to show that the *TNF* rs3093662 polymorphism in the donor impacts the risk of both early and late graft loss after kidney transplantation. To summarize, we demonstrated that the G-allele in the donor nearly doubled the risk of immediate graft loss, while the relative risk of late graft loss increased by 58%. *In vivo* evidence regarding the functional consequences of the *TNF* rs3093662 polymorphism was recently presented, establishing the association of the G-allele with higher serum levels of TNF-α in both healthy and diseased individuals.^22^ Previously, Israni *et al*. also found an association between the *TNF* polymorphism rs3093662 G-allele in the donor and DGF after kidney transplantation.^23^ Although we could not confirm this association (data not shown), the results of Israni and colleagues are in line with our findings that this high-producing variant is detrimental in kidney allografts. Furthermore, in accordance with our study, the G-allele of the *TNF* rs3093662 polymorphism has previously been associated with increased risks of rheumatoid arthritis, adverse events in cancer patients as well as with the susceptibility to and severity of pneumonia.^22,24,25^ Altogether, these studies suggest a pro-inflammatory effect of this high-producing *TNF* polymorphism, thereby increasing the risk for a wide spectrum of immune-mediated diseases.

The clinical use of TNF-α targeted therapeutics has revolutionized the management of inflammatory disorders and autoimmune diseases. In preclinical transplantation studies, TNF-α blockade showed significant protection against renal ischemia-reperfusion injury.^12–14^ The underlying protective mechanisms behind targeting TNF-α include: (i) Modulating immune cell functions, (ii) Inhibiting inflammatory mediators, (iii) Protecting kidney cells from apoptosis, and (iv) Stimulating tissue regeneration.^7,10^ Application of TNF-α antagonists in kidney transplantation would be especially interesting given the extensive clinical experience with this biological. Recently, a meta-analysis found no significant increase in the infection rate among transplantation patients already on TNF-α inhibitors due to inflammatory bowel disease.^26^ However, an observational study, comparing seven renal transplant recipients who resumed their TNF-α inhibitors after transplantation to seven recipients who did not, found that malignancies occurred more frequently.^27^ Etanercept, a fusion protein TNF-α inhibitor, has already been tested in kidney transplantation during hypothermic machine perfusion. However, no differences were seen in renal transplant outcomes between the groups.^28^ Finally, a Phase 2, double-blind, multicenter, randomized controlled trial of 300 deceased donor kidney transplant recipients has just been completed (NCT02495077). In this trial, a mAb targeting TNF-α (Infliximab) was given to recipients prior to transplantation, in addition to current immunosuppressive regimens, with kidney function at 2 years post-transplantation as the primary outcome. The results of this trial are anxiously awaited by the transplant community and will help define the clinical role of TNF-α inhibitors in kidney transplantation.

The present study found that a *TNF* genetic variant (rs3093662 A>G) in the donor, but not in the recipient, associated with the risk of graft loss after kidney transplantation. Our findings indicate that it is not circulating TNF-α from the recipient, but rather the release of TNF-α by the allograft that contributes to kidney transplant failure. These results provide important considerations for anti-TNF-α therapies in kidney transplantation. TNF-α inhibitors that are currently used in clinical practice are either antibodies or soluble TNF-α receptors with distinct pharmacodynamic profiles and side-effects related to systemic exposure. However, if the donor kidney is the site of action, small interfering RNA (siRNA) or antisense oligonucleotides could be a more favorable approach due to better drug penetration into the donor kidney.^29^ The association we found between donor genetics and transplant outcomes suggests that donor pre-treatment with TNF-α inhibitors should also be considered. Interventions within the donor may protect against a proinflammatory reaction after transplantation, and therefore, donor pre-treatment could be a promising strategy to increase graft function and survival.^30,31^ Conversely, donor pre-treatment is not possible until it is known whether other donor organs (i.e., the liver, heart, and lungs) also benefit from targeting TNF-α.

The current study has several limitations that warrant consideration. First, a causality of the observed association cannot be established by this observational study as further work must be done to confirm our results. Second, we only investigated the association of two *TNF* polymorphisms with outcome and did not look at *TNF* haplotypes. Third, because of the lack of plasma samples, we could not examine the differences in TNF-α plasma levels between the *TNF* genotypes. However, important strengths of our study include the prospective design, the large sample size, the long follow-up time, and the clinically meaningful endpoint (i.e., graft loss).

In conclusion, we found that kidney allografts possessing a *TNF* polymorphism, that has been associated with higher TNF-α levels, have a greater risk of immediate and late graft loss after kidney transplantation. Our study adds to the growing body of literature showing that TNF-α blockade in kidney transplantation has the potential to improve transplant outcomes. Following up on this work with clinical trials using TNF-α inhibitors is necessary to determine the ideal setting, including the timing of intervention, in which these biologicals might be effective.

## Data Availability

Data is available upon request.

## Acknowledgment

The authors thank the members of the REGaTTA cohort (REnal GeneTics TrAnsplantation; University Medical Center Groningen, University of Groningen, Groningen, the Netherlands): S. J. L. Bakker, J. van den Born, M. H. de Borst, H. van Goor, J. L. Hillebrands, B. G. Hepkema, G. J. Navis and H. Snieder. The illustrations of Figure 1 were made by Siawosh K. Eskandari.

## Abbreviations

DGF: Delayed graft function
HLA: Human leukocyte antigen
HR: Hazard ratio
OD: Odds ratio
PNF: Primary non-function
mAb: Monoclonal antibody
siRNA: Small interfering RNA
SNP: Single-nucleotide polymorphism
TNF-α: Tumor necrosis factor alpha
*TNF*: Tumor necrosis factor alpha gene

## Supplementary Figures

**Figure S1.**
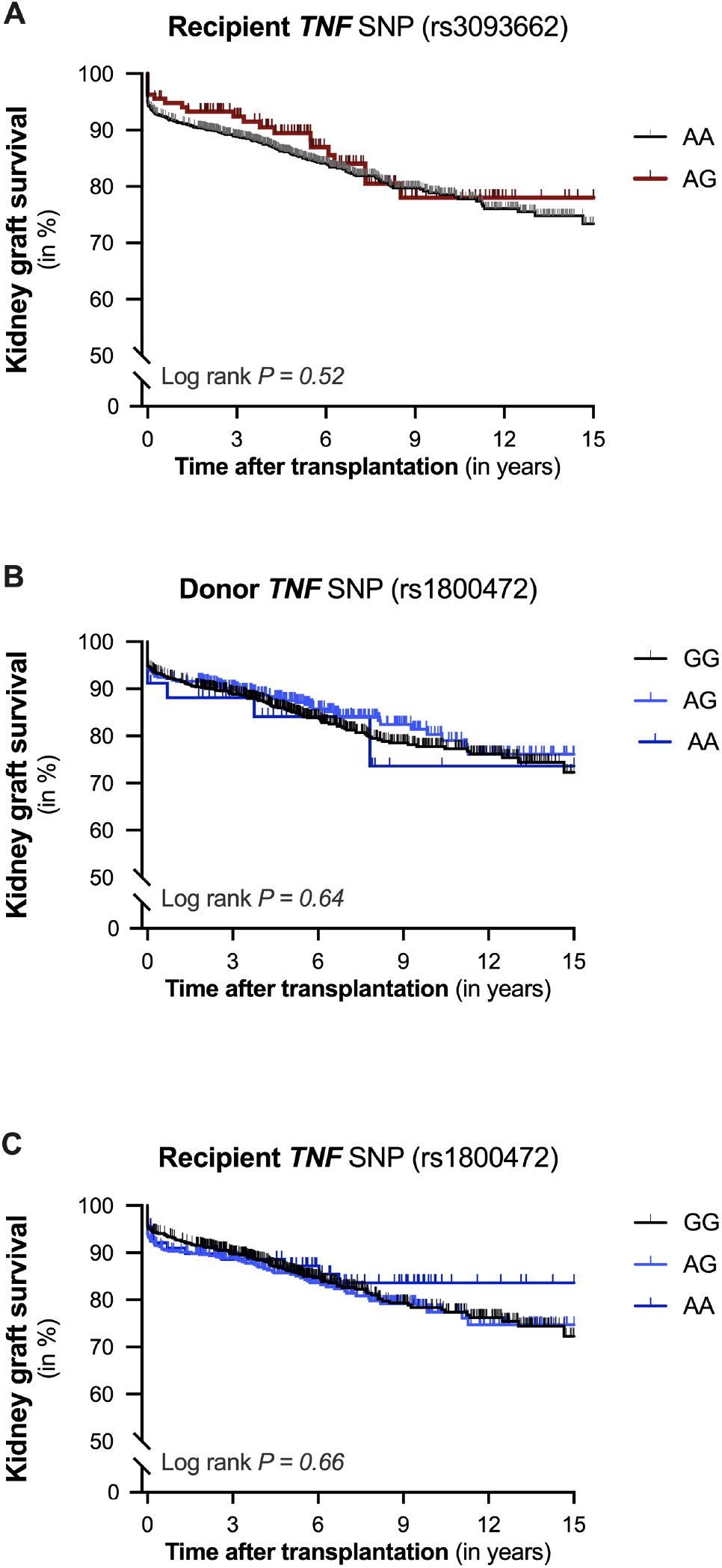
Kaplan-Meier curves for 15-year death-censored graft survival after kidney transplantation according to the presence of TNF variant in the donor and recipient. Cumulative 15-year death-censored kidney graft survival according to the presence of (**A**) the rs3093662 A>G polymorphism in the tumor necrosis factor-alpha gene (*TNF*) in the recipient, and the presence of the rs1800472 G>A polymorphism in *TNF* in the (**B**) donor and (**C**) recipient. The Log-rank test was used to compare the graft loss incidence between the different groups.

## References

1. Wekerle T, Segev D, Lechler R, Oberbauer R. Strategies for long-term preservation of kidney graft function. Lancet. 2017;389(10084):2152–2162. doi:10.1016/S0140-6736(17)31283-7

2. Breyer MD, Susztak K. The next generation of therapeutics for chronic kidney disease. Nat Rev Drug Discov. 2016;15(8):568–588. doi:10.1038/nrd.2016.67

3. Stegall MD, Troy Somerville K, Everly MJ, et al. The importance of drug safety and tolerability in the development of new immunosuppressive therapy for transplant recipients: The Transplant Therapeutics Consortium’s position statement. Am J Transplant. 2019;19(3):625–632. doi:10.1111/ajt.15214

4. Edmonston DL, Roe MT, Block G, et al. Drug Development in Kidney Disease: Proceedings From a Multistakeholder Conference. Am J Kidney Dis. 2020;76(6):842–850. doi:10.1053/J.AJKD.2020.05.026

5. Rudrapal MJ. Khairnar SG. Jadhav A. Drug Repurposing (DR): An Emerging Approach in Drug Discovery. In: Drug Repurposing - Hypothesis, Molecular Aspects and Therapeutic Applications. IntechOpen; 2020. doi:10.5772/intechopen.93193

6. Holdsworth SR, Can PY. Cytokines: Names and numbers you should care about. Clin J Am Soc Nephrol. 2015;10(12):2243–2254. doi:10.2215/CJN.07590714

7. Brenner D, Blaser H, Mak TW. Regulation of tumour necrosis factor signalling: Live or let die. Nat Rev Immunol. 2015;15(6):362–374. doi:10.1038/nri3834

8. Lim H, Lee SH, Lee HT, et al. Structural biology of the TNFα antagonists used in the treatment of rheumatoid arthritis. Int J Mol Sci. 2018;19(3). doi:10.3390/ijms19030768

9. Jang D, Lee A-H, Shin H-Y, et al. The Role of Tumor Necrosis Factor Alpha (TNF-α) in Autoimmune Disease and Current TNF-α Inhibitors in Therapeutics. Int J Mol Sci. 2021;22(5):1–16. doi:10.3390/IJMS22052719

10. Kalliolias GD, Ivashkiv LB. TNF biology, pathogenic mechanisms and emerging therapeutic strategies. Nat Rev Rheumatol. 2016;12(1):49–62. doi:10.1038/nrrheum.2015.169

11. Feldmann M, Pusey CD. Is there a role for TNF-α in anti-neutrophil cytoplasmic antibody-associated vasculitis? Lessons from other chronic inflammatory diseases. J Am Soc Nephrol. 2006;17(5):1243–1252. doi:10.1681/ASN.2005121359

12. Nagata Y, Fujimoto M, Nakamura K, et al. Anti-TNF-α agent infliximab and splenectomy are protective against renal ischemia-reperfusion injury. Transplantation. 2016;100(8):1675–1682. doi:10.1097/TP.0000000000001222

13. Di Paola R, Genovese T, Impellizzeri D, Ahmad A, Cuzzocrea S, Esposito E. The renal injury and inflammation caused by ischemia-reperfusion are reduced by genetic inhibition of TNF-αR1: A comparison with infliximab treatment. Eur J Pharmacol. 2013;700(1-3):134–146. doi:10.1016/j.ejphar.2012.11.066

14. Tasdemir C, Tasdemir S, Vardi N, et al. Protective effect of infliximab on ischemia/reperfusion-induced damage in rat kidney. Ren Fail. 2012;34(9):1144–1149. doi:10.3109/0886022X.2012.717490

15. Moers C, Pol RA, De Borst MH. Tumor necrosis factor a blockade to ameliorate renal ischemia reperfusion injury: Potential implications for kidney transplantation. Transplantation. 2016;100(8):1601–1602. doi:10.1097/TP.0000000000001221

16. King EA, Wade Davis J, Degner JF. Are drug targets with genetic support twice as likely to be approved? Revised estimates of the impact of genetic support for drug mechanisms on the probability of drug approval. PLoS Genet. 2019;15(12):e1008489. doi:10.1371/journal.pgen.1008489

17. Nelson MR, Tipney H, Painter JL, et al. The support of human genetic evidence for approved drug indications. Nat Genet. 2015;47(8):856–860. doi:10.1038/ng.3314

18. Dessing MC, Kers J, Damman J, Navis GJ, Florquin S, Leemans JC. Donor and recipient genetic variants in NLRP3 associate with early acute rejection following kidney transplantation. Sci Rep. 2016;6. doi:10.1038/srep36315

19. Damman J, Daha MR, Leuvenink HG, et al. Association of complement C3 gene variants with renal transplant outcome of deceased cardiac dead donor kidneys. Am J Transplant. 2012;12(3):660–668. doi:10.1111/j.1600-6143.2011.03880.x

20. Damman J, Kok JL, Snieder H, et al. Lectin complement pathway gene profile of the donor and recipient does not influence graft outcome after kidney transplantation. Mol Immunol. 2012;50(1-2):1–8. doi:10.1016/j.molimm.2011.11.009

21. Auton A, Abecasis GR, Altshuler DM, et al. A global reference for human genetic variation. Nature. 2015;526(7571):68–74. doi:10.1038/nature15393

22. Zhang S, Zhan L, Zhu Y, Sun H, Xu X. Tumor Necrosis Factor Alpha Gene Polymorphisms Increase Susceptibility to Adenovirus Infection in Children and Are Correlated with Severity of Adenovirus-Associated Pneumonia. Genet Test Mol Biomarkers. 2020;24(12):761–770. doi:10.1089/gtmb.2020.0122

23. Israni AK, Li N, Cizman BB, et al. Association of Donor Inflammation- and Apoptosis-Related Genotypes and Delayed Allograft Function After Kidney Transplantation. Am J Kidney Dis. 2008;52(2):331–339. doi:10.1053/j.ajkd.2008.05.006

24. Tang R, Sinnwell JP, Li J, Rider DN, de Andrade M, Biernacka JM. Identification of genes and haplotypes that predict rheumatoid arthritis using random forests. BMC Proc. 2009;3(Suppl 7):S68. doi:10.1186/1753-6561-3-s7-s68

25. Kühl T, Behrens S, Jung AY, et al. Validation of inflammatory genetic variants associated with long-term cancer related fatigue in a large breast cancer cohort. Brain Behav Immun. 2018;73:252–260. doi:10.1016/j.bbi.2018.05.009

26. van Meeteren MJW, Hayee B, Inderson A, et al. Safety of anti-TNF treatment in liver transplant recipients: A systematic review and metaanalysis. J Crohn’s Colitis. 2017;11(9):1146–1151. doi:10.1093/ecco-jcc/jjx057

27. Quinn CS, Jorgenson MR, Descourouez JL, Muth BL, Astor BC, Mandelbrot DA. Management of Tumor Necrosis Factor α Inhibitor Therapy After Renal Transplantation: A Comparative Analysis and Associated Outcomes. Ann Pharmacother. 2019;53(3):268–275. doi:10.1177/1060028018802814

28. Diuwe P, Domagala P, Durlik M, Trzebicki J, Chmura A, Kwiatkowski A. The effect of the use of a TNF-alpha inhibitor in hypothermic machine perfusion on kidney function after transplantation. Contemp Clin Trials. 2017;59:44–50. doi:10.1016/j.cct.2017.05.013

29. Gareb B, Otten AT, Frijlink HW, Dijkstra G, Kosterink JGW. Review: Local tumor necrosis factor-α inhibition in inflammatory bowel disease. Pharmaceutics. 2020;12(6):1–31. doi:10.3390/pharmaceutics12060539

30. de Vries DK, Wijermars LGM, Reinders MEJ, Lindeman JHN, Schaapherder AFM. Donor pre-treatment in clinical kidney transplantation: A critical appraisal. Clin Transplant. 2013;27(6):799–808. doi:10.1111/ctr.12261

31. O’Neill S, Oniscu GC. Donor pretreatment and machine perfusion: Current views. Curr Opin Organ Transplant. 2020;25(1):59–65. doi:10.1097/MOT.0000000000000725

